# Salivary Dysbiosis Aligns with an Olfactory–Cognitive Phenotype in Aging

**DOI:** 10.64898/2026.02.12.26346193

**Authors:** Elindi de Coning, Aatmika Barve, Lavinia Alberi, Claire Bertelli, Kevin Richetin

## Abstract

**Background:** Scalable, non-invasive markers for cognitive-decline risk are limited. Olfactory dysfunction is predictive, and oral dysbiosis is mechanistically linked to neurocognitive pathways. Hence, we tested whether pairing smell and global cognition with salivary microbiome profiling yields a targeted, clinically useful signal.

**Methods:** We enrolled 113 Memory Center attendees and community controls. Same-day MMSE, UPSIT, and saliva were obtained for 16S rRNA gene sequencing and cytokine measurement. Unsupervised k-means clustering on standardized MMSE–UPSIT defined two groups of participants: CNN (cognitively normal, normosmia) and CIH (cognitively impaired, hyposmia). Ordination and elastic-net models adjusted for age, sex, BMI, and sequencing depth. Functions were inferred with PICRUSt2 and were integrated with taxa via DIABLO.

**Results:** Overall, the 16S-based microbial community structure was similar between groups, indicating minor compositional shifts. CIH showed enrichment of periodontal anaerobes (*Porphyromonas, Treponema* and *Prevotella*), whereas CNN retained nitrate-reducing commensals (e.g. *Neisseria subflava, Aggregatibacter aphrophilus*). Functional shifts showed mixed consistency with literature, aligning for outer membrane usher proteins and alkyldihydroxy phosphate synthase, but diverging for thiaminase, alpha-glucuronidase, and chemotaxis protein CheX. Most salivary cytokines levels did not differ between groups.

**Conclusions:** This integrated smell, cognition, and saliva workflow delineates an olfactory– cognitive phenotype linked to a targeted, potentially modifiable salivary dysbiosis, periodontal anaerobes vs nitrate-reducers, rather than diffuse salivary inflammatory elevation. This approach may support non-invasive triage and monitoring along the oral– brain axis, pending independent, longitudinal validation.

## Introduction

Cognitive decline is a growing public-health challenge that erodes independence and quality of life in aging populations. Anticipating its onset and trajectory remains difficult because clinical presentations overlap across syndromes and because scalable, non-invasive risk markers are limited. Olfactory dysfunction, especially impaired odor identification, emerges early and predicts subsequent cognitive decline and dementia in both community and clinical cohorts, making brief smell testing an attractive, low-burden tool for risk stratification (Devanand et al., 2014; Wilson et al., 2007).

In parallel, converging epidemiologic and mechanistic evidence implicates the oral microbiome in systemic inflammation and neurocognitive outcomes, positioning an oral– brain axis as a plausible way to link oral ecology to brain function (Bathini et al., 2020; Dominy et al., 2019; Fogelholm et al., 2023; Sadrameli et al., 2020; Yang et al., 2021). The oral cavity hosts one of the body’s most diverse microbial ecosystems, whose composition displays geographical and population-level variation. Location and lifestyle represent significant determinants of community structure, underscoring the need for multi-site, context-aware frameworks (Nasidze et al., 2009; Ruan et al., 2022; Wang et al., 2022).

Disruptions in the oral microbiome contribute to periodontal disease primarily through microbial dysbiosis rather than infection by a single pathogen (Radaic & Kapila, 2021). The “red complex” bacteria, *Porphyromonas gingivalis, Tannerella forsythia*, and *Treponema denticola*, are strongly linked to severe periodontitis. These anaerobes colonize deep subgingival pockets, where they induce chronic local inflammation and tissue destruction (Holt & Ebersole, 2005; Maekawa et al., 2014). In contrast, “orange complex” species such as *Prevotella intermedia, Fusobacterium nucleatum*, and *Campylobacter gracilis* inhabit more superficial gingival sites, serving as bridge organisms that facilitate red complex colonization and biofilm maturation (Carrouel et al., 2016; Hajishengallis & Lamont, 2012). Although initially confined to periodontal pockets, this chronic mucosal colonization can disseminate its effects systemically, as periodontal bacteria and their inflammatory mediators reach the circulation and contribute to extraoral pathology (Hajishengallis, 2014a).

Periodontal pathogens, including *P. gingivalis*, whose virulence is largely mediated by its proteolytic toxins, gingipains, together with *F. nucleatum* and *T. denticola*, have been implicated in Alzheimer’s disease (AD) via chronic inflammation and amyloid-β (Aβ) aggregation (Visentin et al., 2023). Beyond infection-inflammation pathways, nitrate-reducing commensals such as *Neisseria* and *Aggregatibacter* sustain the oral conversion of nitrate to nitrite to nitric oxide (NO) that influences vascular tone and, in older adults, cerebral perfusion (Bescos et al., 2020; Bondonno et al., 2015; Presley et al., 2011). Nitric oxide is considered beneficial to oral and vascular health due to its antimicrobial and vasodilatory properties (Blekkenhorst et al., 2018; Hezel & Weitzberg, 2015a), while further conversion to ammonia can help buffer acidification, limiting caries-promoting biofilm development (Rosier et al., 2020).

However, few clinical studies have integrated olfaction, global cognition, and salivary/oral microbiome profiling within a single, pragmatic framework that could translate into a frontline triage strategy. To test this strategy, the present study combined odor identification (UPSIT) and global cognition (MMSE) with 16S-based salivary microbiome profiling and lightweight inflammatory readouts to determine whether discrete, targeted oral microbial signatures predict a combined olfactory–cognitive phenotype. We demonstrate that participants grouped as cognitively impaired with hyposmia based on MMSE–UPSIT present a salivary dysbiosis, characterized by the enrichment of periodontal anaerobes and a depletion of nitrate reducers, without parallel elevations in common salivary cytokines. While not implying causality, these results highlight a measurable microbial signature that may be leveraged for non-invasive triage, monitoring, or as a therapeutic target in future longitudinal and interventional studies along the oral–brain axis (Bondonno et al., 2015; Presley et al., 2011).

## Methods

### Study design and participants inclusion

We conducted a prospective cross-sectional study at the Cantonal Hospital of Fribourg Memory Clinic (Switzerland). The clinical cohort of 113 subjects included patients who came with cognitive complaints and other community controls from the same catchment area. All procedures (clinical exam, cognitive and olfactory testing, saliva collection) were performed same-day between 09:00–11:00 to minimize circadian/dietary variability. The study protocol was approved by the Cantonal Ethics Committee (CER-VD2016-01627), and written informed consent was obtained from all participants (Bathini et al., 2020).

### Cognitive and olfactory assessment

Global cognition was measured using the Mini-Mental State Examination (MMSE) (Folstein et al., 1975). Olfaction was assessed with the chemosensory University of Pennsylvania Smell Identification Test (UPSIT) using smell strips booklet (Sensonics International) according to the manual guidelines (Doty et al., 1984). Scaled MMSE and UPSIT scores were used to cluster patients using unsupervised k-means clustering. The optimal number of clusters was determined using the gap statistic (*clusGap*) with a maximum of 10 clusters and 50 bootstrap iterations (Tibshirani et al., 2001).

### Saliva collection, storage and processing and storage

Participants were instructed to avoid eating/drinking for at least 2 hours prior to sampling. About ~2 mL of unstimulated whole saliva was collected into sterile tubes. Aliquots were stored at −20 °C (short-term) and −80 °C (long-term) until DNA extraction and cytokine profiling. Genomic DNA was extracted using the Norgen Saliva DNA Isolation Kit (RU45400) according to the manufacturer’s protocol. The V3–V4 region of the 16S rRNA gene was amplified with Illumina-tailed primers. Amplicons were bead-purified, indexed (Nextera XT v2), normalized, pooled with 10% PhiX, and sequenced on an Illumina MiSeq (v3 chemistry, 2×300 bp). A no-template control and a commercial mock community (ZymoBIOMICS) were included.

### Microbiota analysis

Reads were processed with zAMP (Scherz et al., 2024), a DADA2-based workflow for error modeling and ASV inference (Callahan et al., 2016). Taxonomy was assigned against EzBioCloud (Chalita et al., 2024; Yoon et al., 2017). ASV tables were imported into phyloseq (McMurdie & Holmes, 2013) for downstream analysis in R (version 4.4.5). Species with prevalence ≤2 samples were removed to reduce sparsity. Because sequencing depth can drive community structure, depth was retained as a covariate in all multivariate models.

We used multivariate Capscale (Anderson & Willis, 2003; Legendre & Gallagher, 2001) analysis to explore the relationship between microbial communities and environmental or experimental variables. The impact of environmental factors (age, BMI, sequencing depth, sex, and clusters) was systematically examined by iterating through all possible combinations as predictors to assess their influence on the response variable, the Bray-Curtis dissimilarity matrix (Bray & Curtis, 1957) derived from the Hellinger-transformed species table.

To identify taxa associated with each phenotype while acknowledging compositionality, we applied elastic-net generalized linear models (glmnet (Friedman et al., 2010)) to centered log-ratio (CLR)-transformed species abundances, adjusting for age, sex, BMI, and sequencing depth. The glmnet procedure applies a penalty (λ) that shrinks less informative features toward zero to reduce overfitting; the optimal λ was chosen by cross-validation at λ_min, the value that minimized prediction error. Taxa with non-zero coefficients were prioritized as those most robustly separating microbial community between phenotypes, with model stability confirmed by resampling and sensitivity analyses (e.g., excluding low-depth libraries).

### Functional inference and integrative analysis

Metagenome-based functions were inferred with PICRUSt2 based on 16S-reference genomic sequences and gene family (KEGG Orthologs, KO) prediction (Kanehisa & Goto, 2000). KO tables underwent the same framework to isolate functions splitting CNN and CIH after covariate adjustment.

Species and KO matrices were integrated using DIABLO (Singh et al., n.d.) from mixOmics package (Rohart et al., 2017), to identify covarying signatures that maximize discrimination and cross-dataset correlation, correcting for multiple testing (Singh et al., n.d.). Inputs were CLR-transformed and went through tuning (prevalence = 0.3) for number of components and keepX per block to choose by repeated cross-validation to balance stability and parsimony.

Co-variation between microbial species and associated KO was visualized as a network in Cytoscape (3.10.3) (Shannon et al., 2003). Lastly, KOs from the nitrogen metabolism pathway (KO00910) present in the functional matrix were extracted. Group means were calculated per cluster (CNN vs. CIH), and Δ values (CIH mean minus CNN mean) were computed. These were color-mapped onto the KO00910 pathway using Bio.Graphics.KGML_vis (Biopython v1.75) in Python 3.11.11.

### Cytokine Profiling

A 16-plex human inflammation panel (Aimplex) was performed on a BD LSRII flow-cytometer (BD Biosciences). Cytokine expression and inflammatory markers were compared between the two-patient groups. The ROUT method of GraphPad Prism (10.3.0) was used to identify the outliers among the dataset which were removed from visualization in boxplots (**Figure 1D and Supplemental Figure 2**). Figures were generated, and two-sided tests and permutation p-values were performed on GraphPad Prism 10.3.

**Figure 1.**
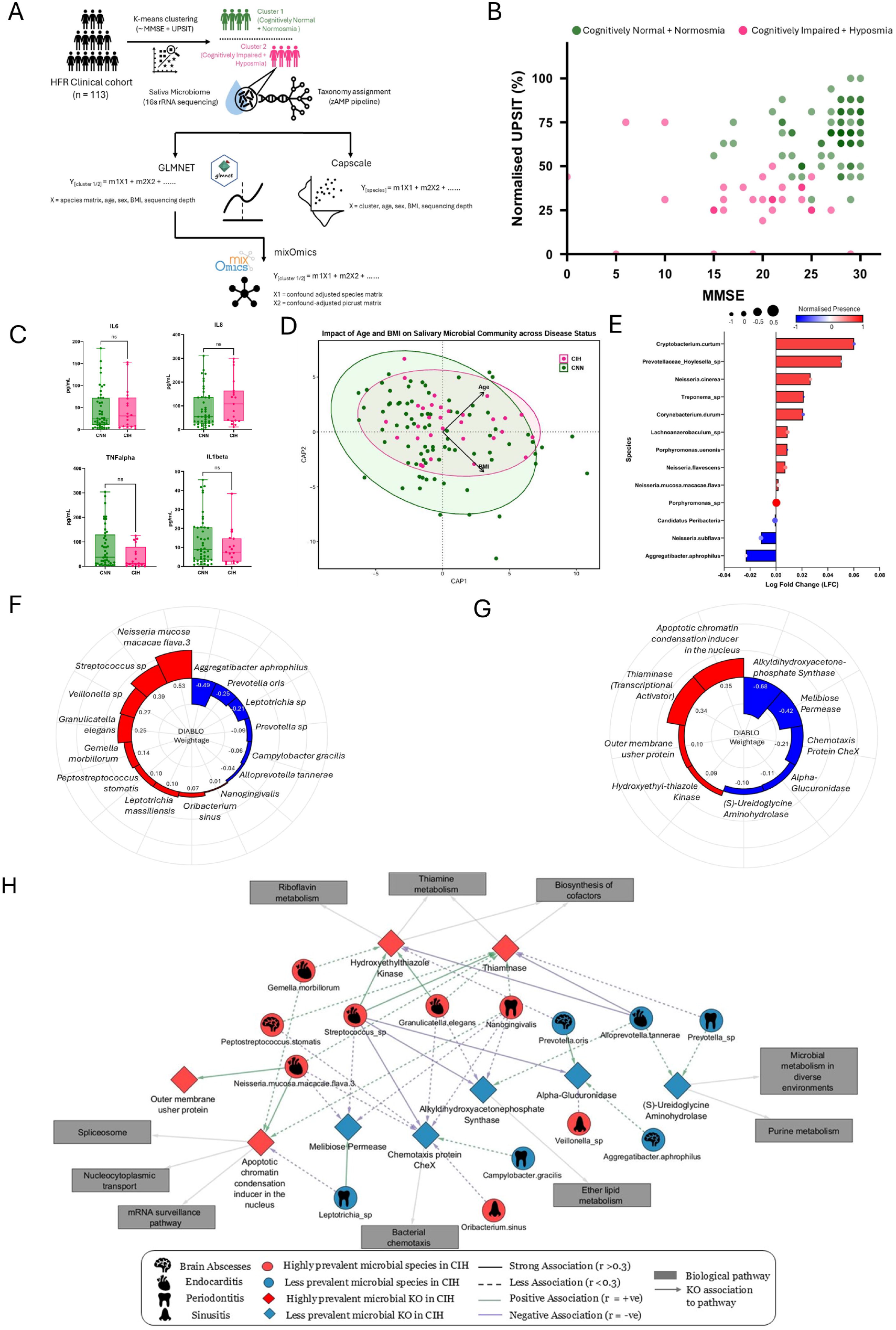
Salivary microbiome and neuro-olfactory status. A) Overview of the saliva microbiome study workflow. B) Scatter plot representing the clinical cohort clustered through the MMSE score and Normalized UPSIT score (%), into Cognitively Normal + Normosmia (CNN) (N = 82; green) and Cognitively Impaired + Hyposmia (CIH) (N = 31; pink). C) Cytokine expression representing no significant difference between CNN and CIH suggesting no infection in the oral cavity; IL6 (pvalue=0.86); IL8 (pvalue=0.41); TNF-alpha (pvalue=0.36); IL1β (pvalue=0.94). D) Beta-diversity of microbial species within two groups represented through CAPscale method showing the effect of BMI and Age (p = 0.17 and 0.07, respectively). Green points are CNN and pink points are CIH. E) Differential abundance of microbial species observed in CIH compared to CNN through the glmnet model. Red bars show species more abundant in CIH and blue bars less abundant species in CIH. The oval size on top of each bar represents normalized prevalence of species. Red and blue ovals present species highly or lowly prevalent in CIH, respectively. F,G) Radial bar plots of species-level (F) and PICRUSt2-predicted KO-level (G) DIABLO loadings (component 1), highlighting covarying microbial taxa/functions that maximize CIH discrimination and cross-dataset correlation. Red bars; taxa/functions more prevalent in CIH (strongly weighted on CIH-discriminant axis), blue bars; less prevalent in CIH (weaker/counter-weighted), tuned at prevalence ≥ 0.3 by repeated cross-validation. Bar lengths reflect relative loading magnitude (no direct biological scale). H) Network of CIH‐associated microbiome features and their functions, pathways, and clinical links. Edge sign/strength between node classes (species, KO) represent DIABLO correlations. Icons represent organ affected/infected by these bacterial species according to literature. Nodes: red circles = highly prevalent microbial species in CIH (species driving CIH-discriminant axis); blue circles = less prevalent species in CIH (species counter-weighted to non-CIH); red diamonds = highly prevalent microbial KEGG orthologs (KOs) in CIH (CIH-weighted KOs); blue diamond = less prevalent KOs in CIH (non-CIH-weighted KOs); grey rectangles = enriched metabolic pathways. Edges represent correlations (coefficient abs(r)): solid lines represent strong association (r > 0.3) and dashed line represent weak association; color of the line represents the sign of correlation, green = positive correlation, purple = negative correlation. The grey arrow represents the associated biological pathways to the KEGG orthologs.

## Results

In this prospective cross-sectional study (Bathini et al., 2020), we enrolled 113 individuals at Cantonal Hospital of Fribourg Memory Clinic (Switzerland), including patients referred to cognitive complaints and community controls from the same catchment, most commonly as relatives sharing households. On the same day, all participants completed MMSE (**Supp Fig 1A**) and UPSIT (**Supp Fig 1B**) testing and provided saliva for 16S rRNA sequencing (**Fig. 1A**). To avoid arbitrary clinical cut-offs and to capture a combined cognitive–olfactory phenotype, we applied k-means clustering on MMSE–UPSIT scores to identify optimal patient clusters (**Supp Fig 1C**). Two phenotypes emerged: individuals cognitively normal with normosmia (CNN) and cognitively impaired with hyposmia (CIH) (**Fig. 1B and Supp Fig 1D**). Given our hypothesis of mucosal inflammation, salivary cytokines were measured. However, most cytokines did not differ between CNN and CIH, arguing against a generalized local inflammatory state in cognitively impaired with hyposmia (**Fig. 1C, Supp Fig 2 (A-L))**.

16S rRNA profiles revealed similar overall community structure between groups, a small fraction (~2.9%) of total compositional variability explained by predictors. Age and BMI vectors, nearly perpendicular with similar lengths, suggest that age and BMI shape distinct microbial niches with comparable influence on beta-diversity. However, their associations with microbial communities were marginally non-significant (p = 0.17 and 0.07, respectively) (**Fig. 1D**). Penalized elastic-net models, adjusted for age, sex, BMI, and sequencing depth, showed enrichment of periodontal anaerobes in CIH (*Porphyromonas, Prevotella, Treponema, and Cryptobacterium curtum*) and retention of nitrate-reducing commensals in CNN (*Neisseria subflava, Aggregatibacter aphrophilus*) (**Fig. 1E–F**).

To explore the potential functional implications of these community shifts, we examined the relative abundance of nitrogen metabolism–related genes (KEGG KOs), focusing on nitrate-reduction modules leading to the production of NO/N□O or ammonia (NH□), which may influence oral - and indirectly systemic - physiology. Overall, several KOs associated with nitrate/nitrite reduction to NO or N□O (K00368, K15864, K04561) were more abundant in CNN samples (blue-coded, with color intensity proportional to abundance; **Supp Fig 3**). KOs involved in conversion to NH□ (K00260, K05601, K00362, K03385) were detected in both groups, although genes related to extracellular nitrate uptake appeared slightly more abundant in CNN. These observations based on relative abundances are descriptive; however, an exploratory differential abundance analysis identified K03385 as enriched in CNN. Genome inspection confirmed the presence of K03385 in *A. aphrophilus*, a species also enriched in CNN (**Supp Fig 4**).

Multi-omics integration with DIABLO condensed taxa–function signals into sparse signatures aligned with CNN/CIH status, organized into coherent modules (**Fig. 1H**). Associations persisted after adjustments for age, sequencing depth, prevalence filters, and alternative initializations. Inferred functional changes included increases in prevalence of outer membrane usher proteins and decreases in alkyldihydroxy phosphate synthase. Network analysis detected elevated *Streptococcus sp*., *Veillonella sp*., *Peptostreptococcus stomatis*, and *Gemella morbillorum* in CIH, alongside higher *Prevotella* species and lower *Alloprevotella tannerae* in CNN.

## Discussion

We propose a pragmatic “smell - cognition - saliva” workflow in which an unfavorable olfactory–cognitive phenotype co-occurs with a targeted salivary dysbiosis, characterized by the enrichment of periodontal anaerobes and the relative depletion of nitrate-reducing commensals, without parallel increases in most salivary cytokines.

These findings support a vasculo-inflammatory model along the oral–brain axis, in which shifts from nitrate-reducing commensals toward periodontal anaerobes may promote systemic inflammation and impair vascular and neuronal function. Notably, the periodontal pathogens enriched in CIH include key members of the red complex, *P. gingivalis* and *T. denticola* (Mohanty et al., 2019), as well as *Prevotella* species belonging to the orange complex (Könönen et al., 2022a) alongside the emerging periodontal taxa, *Cryptobacterium curtum* (Vidya Hiranmayi et al., 2017). These shifts in bacterial communities are consistent with oral dysbiosis patterns linked to chronic inflammatory signaling, disruption of barrier integrity, and neuroinflammatory cascades that can contribute to cognitive decline.

In contrast, the retention of nitrate-reducing taxa such as *N. subflava* and *A. aphrophilus* in CNN suggests preservation of an oral bacterial community that supports vascular nitric oxide bioavailability and healthier cognitive trajectories (Hezel & Weitzberg, 2015b). Indeed, the presence of nitrite reductase (KO03385) in *A. aphrophilus* indicates a capacity for nitrite-to-ammonia conversion, which can alkalinize the oral environment and reduce caries risk by limiting acid-driven demineralization (Hajishengallis, 2014b).

Oral nitrate-reducing communities were suggested to mediate the enterosalivary nitrate to nitrite to NO pathway, influencing endothelial function, cerebral blood flow, and cognitive performance. Diets rich in inorganic nitrate have been shown to favor *Neisseria*-dominated consortia and improve NO-related vascular markers, supporting the idea that loss of these commensals in CIH could exacerbate NO deficiency and neurovascular risk (Vanhatalo et al., 2021). These findings suggest an active nitrogen-metabolism profile in CNN that may simultaneously stabilize oral pH, favor nitrate reduction and controlled ammonia production that may help maintain both oral homeostasis and brain health.

Network analysis further reinforced periodontal associations for *Streptococcus sp*. (Dhotre et al., 2018), *Veillonella sp*. (Mashima & Nakazawa, 2013), *Peptostreptococcus* and *G. morbillorum* (Gomes et al., 2008). Moreover, *Streptococcus* and *Veillonella* were previously linked to cognitive impairment (Da et al., 2023; Wei et al., 2023). We observed unexpected patterns for *Prevotella* species and *Alloprevotella tannerae*, which are well-established periodontal pathogens (Könönen et al., 2022b) but reduced in our CIH group. Notably, *Prevotella oris* was increased in CNN, a finding that aligns with Alzheimer’s disease studies reporting a negative association of *P. oris* with APOE4 carrier status in humans (L’Heureux et al., 2025).

These targeted microbial shifts observed, despite overall community similarity and minimal cytokine differences, indicate that phenotype-specific dysbiosis, rather than generalized inflammation, underlies the CNN/CIH distinction, supporting the need for longitudinal validation. This success underscores the strength of our cognition-olfactory clustering framework, which identified literature-consistent taxa (including two of three red complex species, *Treponema* and *Porphyromonas*) and functions without relying on clinical cutoffs, establishing robust triage for oral-brain axis research. Methodologically, our avoidance of hard cut-points through same-day MMSE–UPSIT clustering, followed by constrained ordination, penalized models, and multi-block DIABLO integration, yielded statistically parsimonious and clinically portable signatures.

The workflow proposed here fits clinical realities. Older adults with MCI have fewer ambulatory visits for chronic conditions, such as hypertension, increasing the likelihood that risk factors are missing. Compressing screening into one appointment (e.g., UPSIT, MMSE, saliva) reduces attrition and costs (Springer et al., 2025). At a systems level, blood-derived peripheral windows, such as clonal hematopoiesis (CHIP), demonstrate that measurable peripheral states can inform central risk. Indeed, larger CHIP clones (VAF ≥ 8%) were inversely associated with probable dementia in WHIMS (Jakubek et al., 2025). By analogy, saliva offers a practical, non-invasive sample that can be embedded in front-line settings to capture brain-relevant biology alongside clinical screening.

## Conclusion

Clinically, combining the MMSE, UPSIT, and saliva may offer a practical, non-invasive triage strategy that could contribute to early risk stratification along the oral–brain axis, upstream of costly investigations. Indeed, the CIH subgroup (cognitive impairment and hyposmia) is linked to a measurable, targeted salivary dysbiosis rather than cytokine elevations. Without inferring causality, the pattern highlights a potentially modifiable oral microbe ecology (nitrate-reducing commensals) and suggests testable avenues for interventions (periodontal care, oral-hygiene optimization, dietary nitrate) with objective monitoring of the taxa/function signature. The generalizability of these signatures now requires evaluation in large, multi-site cohorts across geographical regions and populations, to establish robustness, sensitivity and diagnostic accuracy.

Aligning an olfactory–cognitive phenotype with a functional salivary dysbiosis delineates a biologically coherent and potentially clinically actionable signal. Future studies, including shotgun metagenomics, anchoring to periodontal status and A/T/N biomarkers, reversibility trials, and deployment as a same-day workflow will test the prognostic value, while opening routes to scalable, non-invasive prevention strategies.

## Acknowledgement

This work was supported as a part of Lawson Foundation, MCM Foundation, CHUV foundation, Eurostars 3 CoD 07-CUP H93B24000210003 and Eurostars 7 ReMemo E!7381, as well as NCCR Microbiomes, a National Centre of Competence in Research, funded by the Swiss National Science Foundation [grants number 180575 and 225148] and Donase Foundation to CBL supporting E. de Coning salary.

## Data availability

The data for this study have been deposited in the European Nucleotide Archive (ENA) at EMBL-EBI under accession number PRJEB108211.

## Supplemental Figure Legends

**Supplemental Figure 1.**
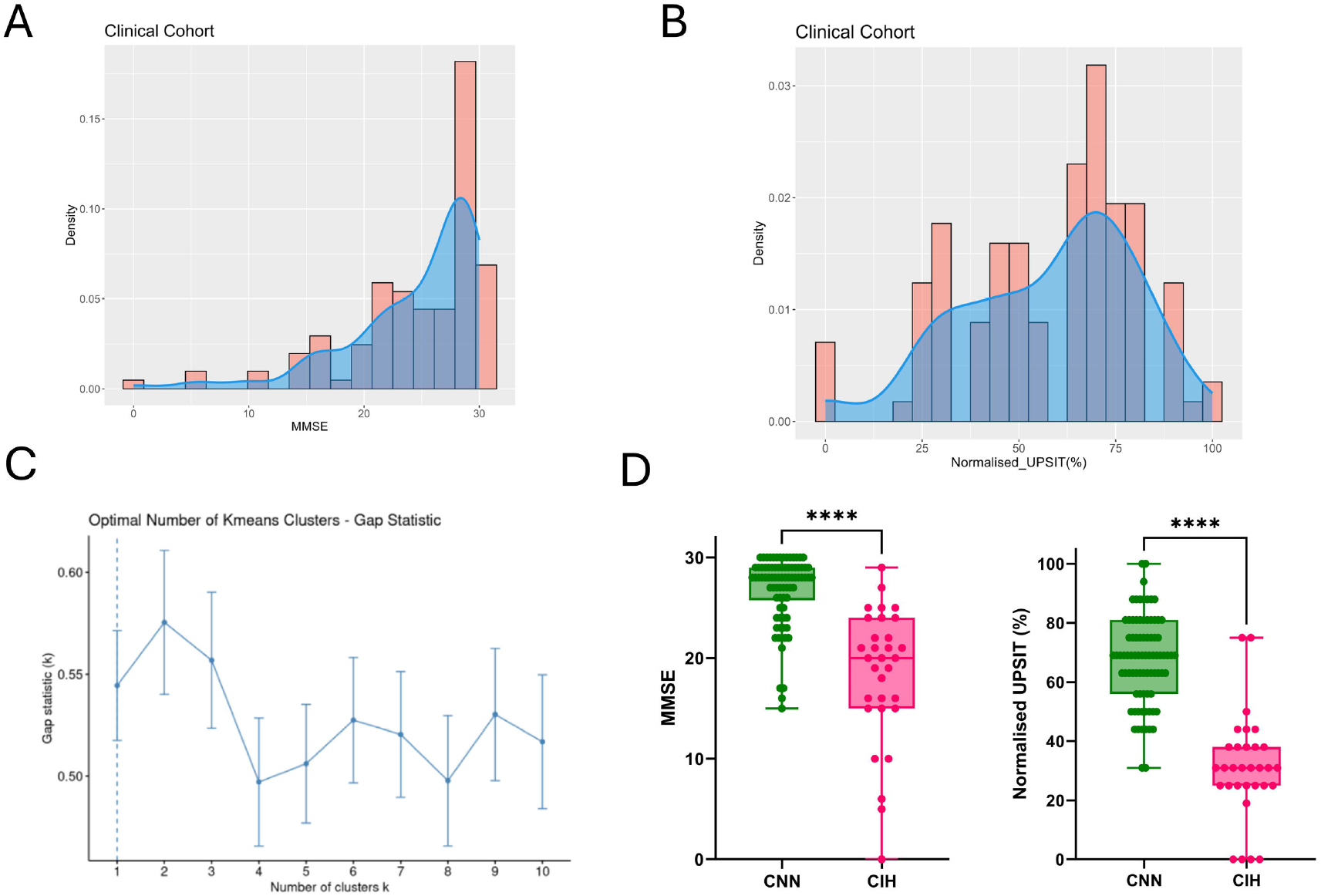
Cognitive and olfactory stratification of the clinical cohort A) Histogram representing the population density of the clinical cohort with the scores for Neuropsychological test MMSE (0-30) and B) for Normalized UPSIT (0%-100%). C) Scree plot representing the Gap-statistics found for different number of k clusters in the clinical cohort. Optimal number of clusters identified using unsupervised k-means clustering was 2 with highest gap statistic value. D) Boxplot representing the difference between CNN and CIH for MMSE scores (pvalue<0.0001) and UPSIT scores (pvalue<0.0001).

**Supplemental Figure 2.**
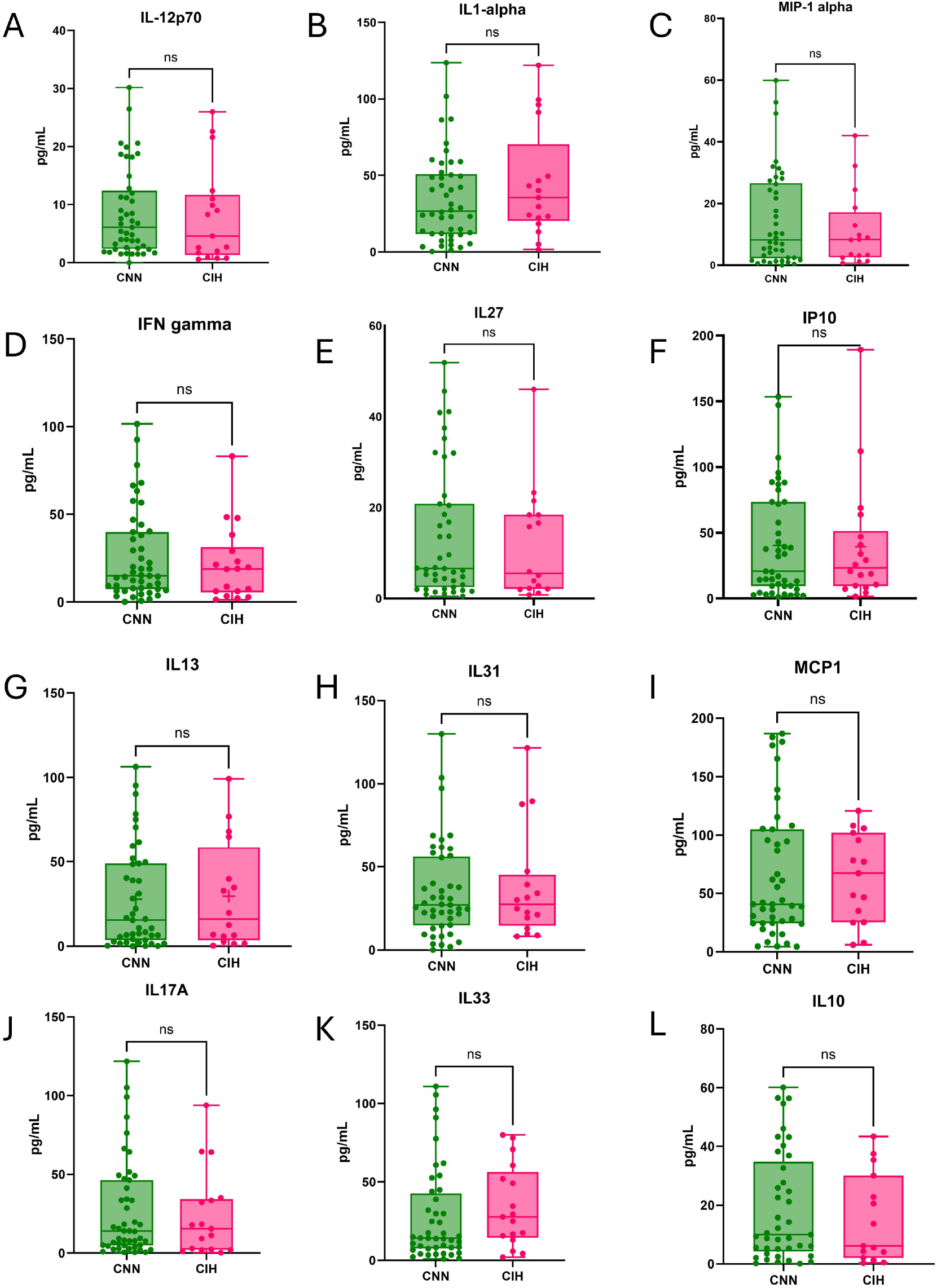
Expression (pg/ml) of different cytokines measured in the immunoassay in the clinical cohort. Boxplot represents the difference between CNN and CIH for A) IL-12p70 (pvalue= 0.9841); B) IL-1 alpha (pvalue= 0.2849); C) MIP-1 alpha (pvalue=0.9599); D) INF gamma (pvalue=0.8998); E) IL27 (pvalue=0.9502); F) IP10 (pvalue=0.9264); G) IL13 (pvalue=0.7661); H) IL31 (pvalue=0.4545); I) MCP1 (pvalue=0.3059); J) IL17A (pvalue=0.9701); K) IL33 (pvalue=0.1088); L) IL10 (pvalue=0.9520).

**Supplemental Figure 3.**
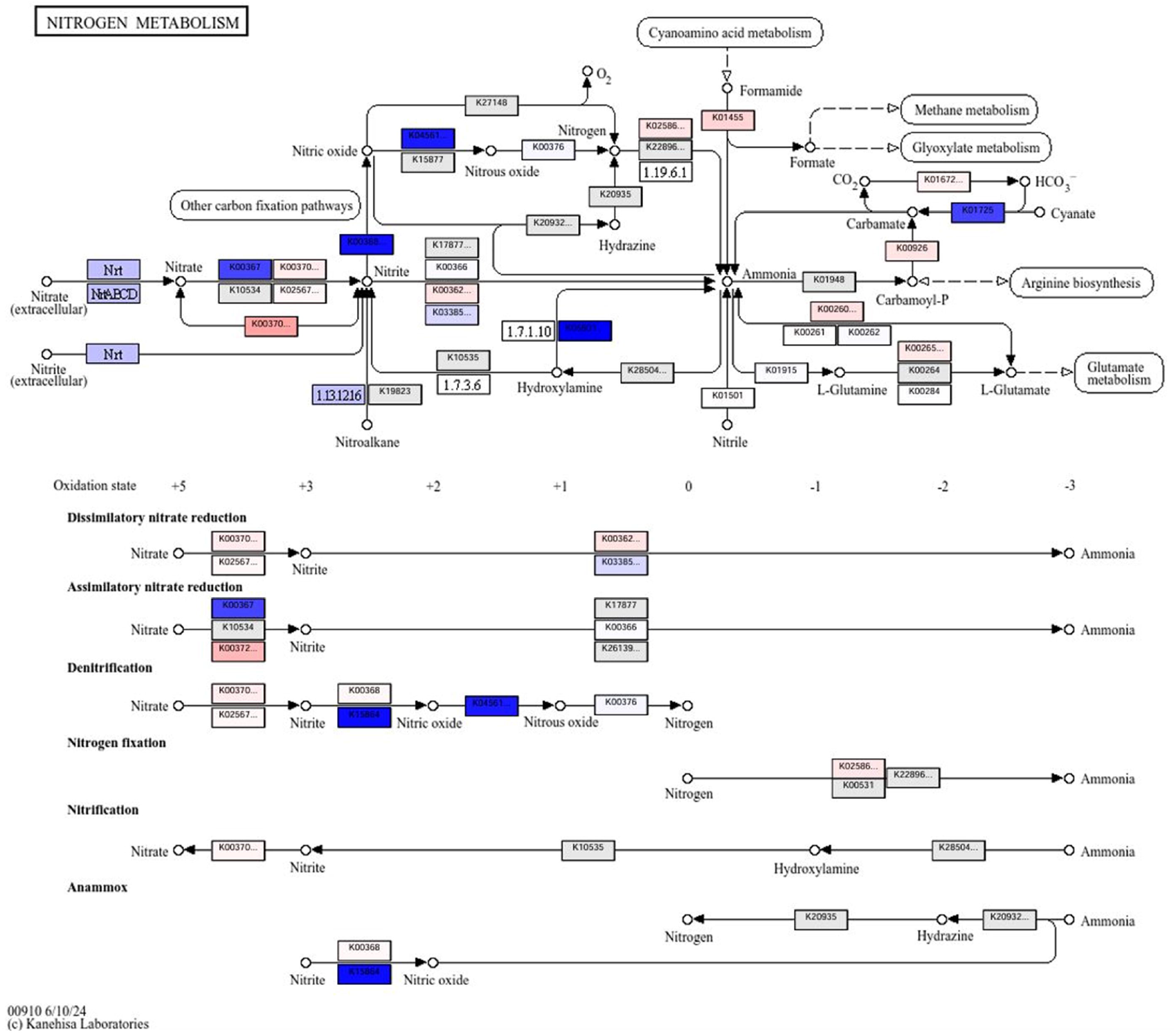
KEGG pathway for Nitrogen Metabolism (KO00910). Only KOs detected in the functional profile are shown. For each KO, group means were calculated per cluster (CNN vs CIH), and Δ values (CIH mean − CNN mean) are displayed. Blue shades indicate lower abundance in CIH (negative Δ), and red shades indicate higher abundance in CIH (positive Δ) along a continuous color spectrum.

**Supplemental Figure 4.**
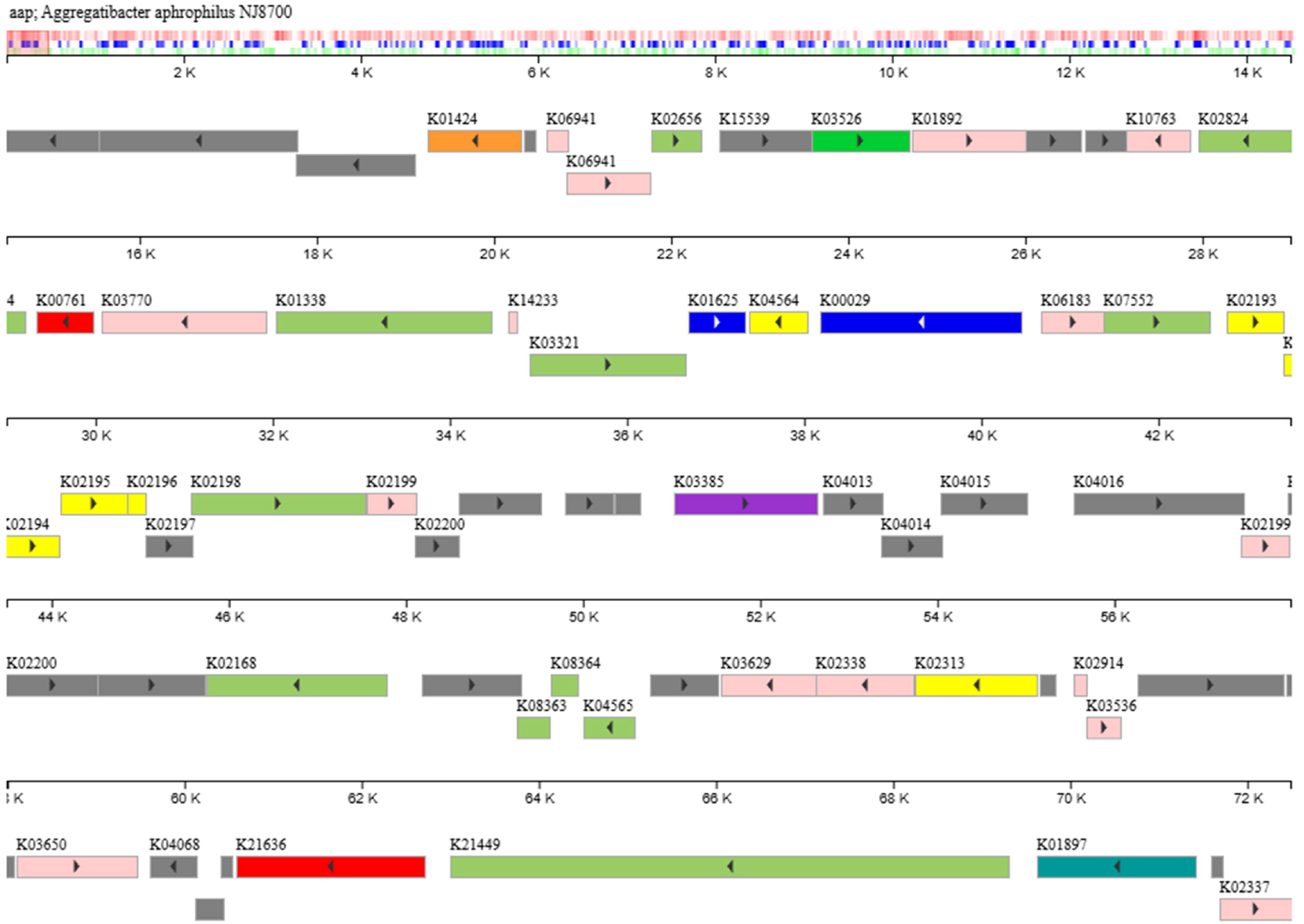
KEGG genome browser for *Aggregatibacter aphrophilus*, confirming the presence of K03385 (nitrate-to-ammonia reduction), which was significantly enriched in CNN samples.

## Notes

### Competing Interest Statement

The authors have declared no competing interest.

### Author Declarations

The study protocol was approved by the Cantonal Ethics Committee (CER-VD2016-01627), and written informed consent was obtained from all participants (Bathini et al., 2020).

